# Atrial fibrillation is associated with decreased claudin-5 in cardiomyocyte

**DOI:** 10.1101/2023.07.11.23292531

**Authors:** Baihe Chen, Haiqiong Liu, Miao Wang, Xianbao Wang, Yuanzhou Wu, Masafumi Kitakaze, Jin Kyung Kim, Yiyang Wang, Tao Luo

## Abstract

**Background:** Although it is critically important to understand the underlying molecular and electrophysiological changes that predispose to the induction and maintenance of atrial fibrillation (AF), the underlying mechanism of AF is still poorly defined. AF is characterized as the electrophysiological and membrane integrity abnormality of the atrial cells, and claudin-5 (Cldn5), a tight junction protein, may be involved in the pathophysiology of AF, however, the role of Cldn5 in AF is unknown.

**Methods:** Left atrial appendages from the enlarged left atrium were obtained from AF patients undergoing modified radiofrequency ablation maze procedure and normal left atrial appendages were obtained from non-AF donors. Western blot, immunofluorescence, transmission electron microscope (TEM), and proteomics analysis were performed to screen the specific protein expression and signal pathway changes in AF heart tissue vs. non-AF heart tissue. In addition, Cldn5 shRNA or siRNA adeno-associated virus (AAV) were then injected into the mouse left ventricle or added into HL1 cells respectively to knockdown claudin-5 in cardiomyocytes to observe whether the change of Cldn5 influences electrophysiology and affects those protein expressions stem from the proteomic analysis. Mitochondrial density and membrane potential were also measured by Mito tracker staining and JC-1 staining under the confocal microscope *in vitro*.

**Results:** The protein level of claudin-5 was significantly decreased in cardiomyocytes from the left atrium of AF patients compared to non-AF donors. Proteomics analysis showed that 83 proteins were downregulated and 102 proteins were upregulated in the left atrial appendage of AF patients. Among them, CACNA2D2, CACNB2, MYL2 and MAP6 were dramatically downregulated. KEGG pathway analysis showed these changes would lead to hypertrophic and/or dilated cardiomyopathy. Cldn5 shRNA AAV infection induced-Cldn5 deficiency caused severe cardiac atrophy and arrhythmias in mice. The decreases in both mitochondrial numbers and mitochondrial membrane potential (MMP) were also observed in vitro after Cldn5 knockdown by siRNA. Finally, western blot analysis confirmed the protein level of CACNA2D2, CACNB2, MYL2 and MAP6 were downregulated after Cldn5 knockdown *in vivo* and *in vitro*.

**Conclusions:** We demonstrated for the first time the deficiency of Cldn5 in cardiomyocytes in the left atrium of AF patients. The mechanism of AF might be associated with Cldn5 deficiency-associated downregulation of CACNA2D2, CACNB2, MYL2 and MAP6, and mitochondrial dysfunction in cardiomyocytes.

**Clinical Perspective:** *What Is New?:*

1. This is the first study to find the decreased expression of claudin-5 (Cldn5) with prominent muscle atrophy in the left atrial appendage of atrial fibrillation (AF) patients.
2. Knockdown of Cldn5 in the left ventricle via shRNA adeno-associated virus (AAV) infection caused myocardial atrophy and arrhythmia including ST elevation, replacement of P-waves with f-waves, and absence of P-waves prior to QRS.
3. The protein levels of CACNA2D2, CACNB2, MYL2 and MAP6 were significantly downregulated after Cldn5 deficiency.

*What Are the Clinical Implications?:* The present findings may improve our understanding of the role of Cldn5 in the pathophysiology of AF and provide a new therapeutic target for preventing AF.

## Introduction

Atrial fibrillation (AF) is the most common arrhythmia encountered in clinics.^1^ Disorganization of electrical impulses in the heart causes a rapid and irregular heart rhythm and thus AF, which leads to substantial morbidity, mortality, and socioeconomic burden worldwide.^2, 3^ Pathogenic mutations of numerous genes have been causally linked to AF, of which the majority encode ion channels, cardiac structural proteins, and gap junction channels.^4, 5^ Gap and tight junctions are the major intercellular junctions that play important roles in cellular communication and structural integrity. In the heart, gap junctions comprised of connexins (Cxs) form intercellular conduits responsible for electrical coupling and rapid coordinated action potential propagation between adjacent cardiomyocytes. Despite accumulating clinical evidence showing that gap junction protein Cx40 and Cx43 abnormalities are associated with AF,^6–9^ there are few investigations for the effects of another cell-cell contact, tight junctions, on electrophysiological properties in human and murine hearts.

Claudin-5 (Cldn5) is a transmembrane tight junction protein that controls endothelial and epithelial permeabilities.^10^ Numerous investigations about the role of Cldn5 have been focused on the blood-brain barrier in the last decades. Interestingly, clinical studies indicate that the level of Cldn5 is reduced in human failing hearts.^11, 12^ Our previous study demonstrated that Cldn5 was expressed in human and murine cardiomyocytes and regulated mitochondrial dynamics by promoting mitochondrial fusion and inhibiting mitochondrial fission, which exerted its protective effect against ischemia insult.^13^ However, the contribution of Cldn5 to cardiac electrophysiology remains poorly defined.

In the current study, we clinically investigated the association of Cldn5 expression with myocardial electrophysiological, morphological, and molecular characteristics in the left atrial appendages of AF patients. Using the Cldn5 shRNA AAV approach, the relationship between Cldn5 deficiency and cardiac electro-structural remodeling was evaluated in mouse heart. The proteomic study was used to assess the protein profile in human left atrial appendages, and western blot was used to confirm the changes of candidate proteins involved in myocardial dilation and atrophy signaling pathways in AF patients’ myocardium and in Cldn5 deficient murine hearts. Finally, cell culture was used to assess Cldn5 knockdown on the mitochondrial density and mitochondrial membrane potential by using Cldn5 siRNA in HL-1 cells. We show that Cldn5 is critical for maintaining normal cardiac morphology and electrophysiology, and downregulation of Cldn5 contributes to cardio-electric disturbance and myocyte atrophy as well as myocardial dilation, which may underly the pathogenesis of AF and lead to novel ways to treat arrhythmias in patients.

## Methods

All procedures were performed in accordance with our institutional guidelines for animal research that conforms to the Guide for the Care and Use of Laboratory Animals, and this study was approved by the Ethical Committee of Zunyi Medical University, Zhuhai Campus. Mice were kept in standard housing conditions with a light/dark cycle of 12 h and free access to food and water.

### Human heart sample isolation

With the approval from a local human research ethics review, left atrial appendages were obtained from patients with severe AF accompanied by rheumatic mitral valve diseases undergoing modified radiofrequency ablation maze procedure or normal control from organ donors that were not used for heart transplantation but had no history of AF and major cardiovascular diseases. Then the tissues were used for western blot, immunofluorescence, and proteomic analysis.

### Echocardiography and Electrocardiography

The AF patients were diagnosed by electrocardiography, and echocardiography was adopted to observe the heart morphology. Cardiac function was evaluated in mice after 4 weeks of Cldn5 shRNA AAV intramyocardial injection by echocardiography and electrocardiography. For echocardiography, using a Vivo-3300 ultrasonic system (VisualSonics, Toronto, ON, Canada) equipped with a 30 MHz high-resolution probe. Mice were anesthetized with inhalational isoflurane at a concentration of 1.5% and two-dimensional parasternal long-axis images of the left ventricle (LV) were obtained at the level of the papillary muscles. The LV anterior wall diameter at diastole (LVAWd) was measured. For electrocardiography, mouse electrocardiography of Techman signal collection and analysis system (BL-420I) were used to track arrhythmias and ventricular premature beat (VPB).

### Adeno-associated virus (AAV)-mediated Cldn5 knockout

C57BL/6 male mice (aged 8–12 weeks, weighing 20–25 g) were anesthetized with a mixture of xylazine (5 mg/kg, intraperitoneal) and ketamine (100 mg/kg, intraperitoneal). Cldn5 shRNA adeno-associated virus (AAV) was injected to the left ventricle by 3-5 points. All mice were under investigation for 4 weeks after Cldn5 shRNA injection to make sure the Cldn5 gene in the myocardium have been successfully knockdown.

### Tandem mass tagging proteomics analysis

The primary experimental procedures for tandem mass tagging (TMT) proteomics analysis include whole-proteome preparation, trypsin digestion, TMT labeling, high-performance pressure liquid chromatography fractionation, LC- MS/MS analysis and data analysis. Raw data were analyzed using GO Annotation (http://www.ebi.ac.uk/GOA/), Domain Annotation (InterProScan) and KEGG Pathway Annotation (KEGG online service tools KAAS mapper).

The protein-protein interaction networks stemming from a computational prediction were analyzed in STRING (https://string-db.org/).

The resulting MS/MS data were processed using Proteome Discoverer search engine (V2.4.1.15) against Homo_Sapiens_9606_PR_20210721.fasta (78,120 sequences) concatenated with reverse decoy database. The parameters were set as follows: (1) trypsin (full) was specified as the cleavage enzyme; (2) two missed cleavages were allowed; (3) the minimum peptide length was six amino acids; (4) the maximum number of modifications per peptide was 3; (5) the mass tolerance for precursor ions was 10 ppm in the first search; (6) fragment ion mass tolerance was 0.02 Da; (7) Carbamidomethylation on cysteine was fixed modification; (8) oxidation on methionine and N-terminal acetylation were variable modification; and (9) false discovery rate was adjusted to 1%. Student’s t-test was used to evaluate the significant differences. The proteins with a fold change of ≥1.50 or ≤1/1.5 and p-value < 0.05 were considered differentially expressed proteins (DEPs).

### Immunofluorescence and Wheat Germ Agglutinin (WGA) staining

Cldn5 was localized by double-label immunofluorescence confocal microscopy, and their overlap was quantified. Briefly, the human and mice heart tissues were fixed in 4% paraformaldehyde, embedded in paraffin, and sections (3–5μm thickness) were stained with primary antibodies for Cldn5-Alexa Fluor 488 (Invitrogen, Catalog#352588), CD31 (Proteintech, Catalog#11265-1-AP) or Tomm20 (Abcam, ab186734) for 1h at room temperature. Then Fluorescein donkey anti-rabbit IgG Alexa Fluor 594 was incubated for 30 min in the dark before confocal microscopic observation. In addition, the cross-sectional areas of the cardiomyocytes in human and mouse myocardium were observed by fluorescein-conjugated wheat germ agglutinin (WGA; 5 µg/mL, 25530, AAT Bioquest, USA) staining and evaluated by calculating the single myocyte cross-sectional areas measured by ImageJ software (National Institutes of Health, USA).

### Cell culture, mitochondrial number and mitochondrial membrane potential (Δψm) analysis

Mouse Cldn5 siRNA (RiboBio, siG2302130920115720) was transfected by lipo 2000 to knock down the expression of Cldn5 in HL-1 cells. Then the mitochondria were imaged by Mitotracker Green FM (Invitrogen, M7514) and the mitochondrial membrane potential was examined by a JC-1 staining kit (Beyotime, C2006). The intensity of the fluorescence was measured by Image J.

### Western Blot Analysis

Proteins were obtained from whole hearts or HL-1 cells. Samples containing equal amounts of protein (10 µg) were separated by 10% SDS-PAGE and transferred onto PVDF membranes. The membranes were blocked with 5% skimmed milk at room temperature for 2 h and then incubated overnight at 4°C with the primary antibody. The following antibodies were used for the Western blotting analysis: Cldn5 (Abcam, ab172968), CACNA2D2 (Abcam, ab173293), CACNB2 (Abcam, ab253193), MYL2 (Affinity, DF7911), MAP6 (Santa Cruz, sc-137036). After being incubated with goat anti-rabbit or goat anti-mouse secondary antibody for 1 h at room temperature, the blots were detected and quantified by densitometry using the Image J Analysis software.

### Transmission Electron Microscopy

Human and mouse heart samples were fixed in 2% glutaraldehyde, phosphate-buffered to pH 7.4 and post-fixed with 1% OsO4. After dehydration, the samples were incubated in propylene oxide followed by embedding in a mixture of Epon 812 and Araldite. Ultrathin sections obtained by an Em UC7 Ultramicrotome (Leica) were collected on TEM nickel grids and analyzed using a TEM (JEM-1400 Plus, JEOL) at 100 kV.

### Statistical analysis

All experiments were expressed as the mean ± SEM and *p* < 0.05 was considered to indicate statistical significance. The paired *t*-test was used for comparisons between the two groups. One-way ANOVA with post hoc analysis by the Fisher exact probability test was employed for multiple comparisons. All analyses were performed using SPSS 13.0 software (SPSS Inc., Chicago, IL).

## Results

### Claudin-5 is downregulated in cardiomyocytes from Af patients’ left atrial appendages

Diagnosis of AF is confirmed by a 12-lead electrocardiogram (ECG) and an echocardiographic observation in an apical four-chamber view. The representative images of ECG and echocardiography were presented in Figure 1A in an AF patient. A significantly enlarged left atrium (LA) was observed (red dashed box in Figure 1A). To mitigate the risk of stroke, we adopted a modified radiofrequency ablation maze procedure to ablate AF triggers and modify AF substrates (Figure 1B, right panel), which is proven more efficient than the catheter-based introduction of lesions (Figure 1B, left panel). The excised LA appendages were then used for western blot and immunofluorescence (IF) to observe the Cldn5 expression and cardiomyocyte morphology. The protein level of Cldn5 was significantly decreased in the AF group (Figure 1C). The IF images further showed the localization of Cldn5 on mitochondria and confirmed the decreased expression of Cldn5 in cardiomyocytes (Figures 1D and 1E). Left atrial appendage sections stained with FITC-WGA also showed the decreased cross-section of cardiomyocytes in the AF group when compared with non-AF.

**Fig. 1.**
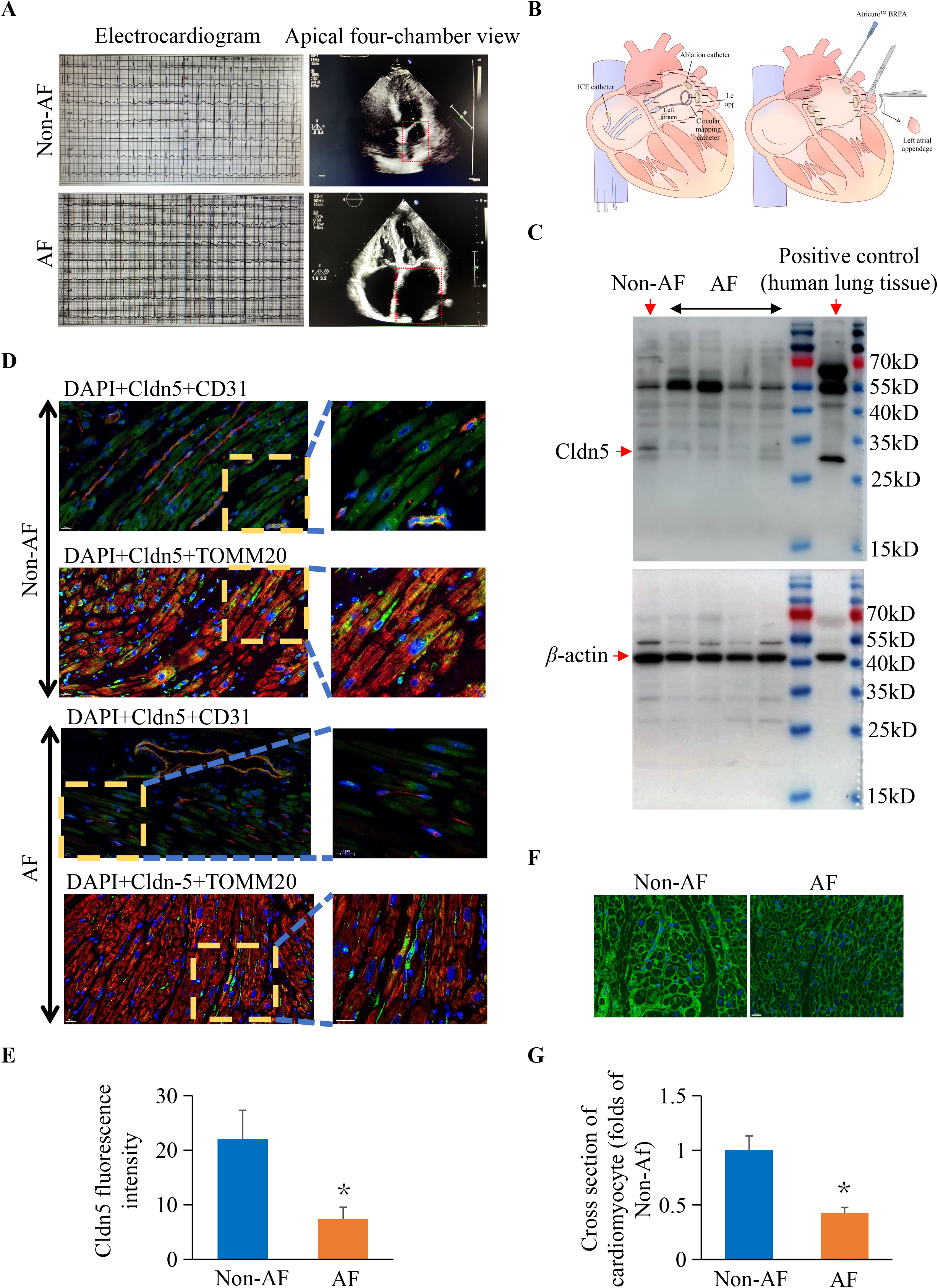
Myocardial dilation, myocyte atrophy and Cldn5 reduction in the left atrial appendages of AF patients. (A) Exemplar images of electrocardiogram (ECG) and apical four-chamber view of the heart in AF patient and control. Red box indicates left atrium (LA). (B) Left appendages of the LA was excised during the modified radiofrequency ablation maze procedure indicated in the right panel. Left panel indicates the traditional percutaneous radiofrequency ablation for AF. (C) Representative western blot of Cldn5 in left appendages of AF patients. Human lung tissue is used as positive control. (D) Exemplar left atrial appendage section stained with anti-Cldn5, CD31 and TOMM20 antibodies. Anti-Cldn5 primary antibody is Alexa Fluor 488 conjugated (Green) while CD31 and TOMM20 antibodies is visualized by Fluorescein donkey anti-rabbit IgG Alexa Fluor 594 (Red). DAPI is used to stain nucleus. Scale bar, 20 µm. (E) Quantification of immunofluorescent staining of Cldn5 in cardiomyocytes. (F) Exemplar left atrial appendage section stained with WGA for assessing cardiomyocyte’s cross section. Scale bar, 20 µm. (G) Quantification of cross-section of cardiomyocytes. * *P* < 0.05 vs. Non-AF; n = 5.

### Claudin-5 knockdown causes cardiac atrophy and disrupts cardiac rhythm in mice

In-vivo studies in C57BL6 mice were performed to evaluate the influence of cardiac Cldn5 on cardiac morphology and electrophysiology. Cldn5 shRNA AAV was injected in the murine left ventricular myocardium for 4 weeks to knock down the Cldn5 as shown in Figure 2A. Western blot showed AAV infection caused a 69.5% reduction of Cldn5 in the LV (Figures 2B and 2C). The thickness of the LV anterior wall was also reduced by 45% in the shRNA AAV infection group (Figures 2D and 2E). Cardiac conduction was assessed by II-lead ECG specialized for murine, which detected the arrhythmia including ST elevation, replacement of P-waves with F-waves and absence of P-waves prior to QRS (Figure 2F). Additionally, LV sections stained with FITC-WGA showed a decreased cross-section of cardiomyocytes in the shRNA infection group compared to the control (Figures 2G and 2H).

**Fig. 2.**
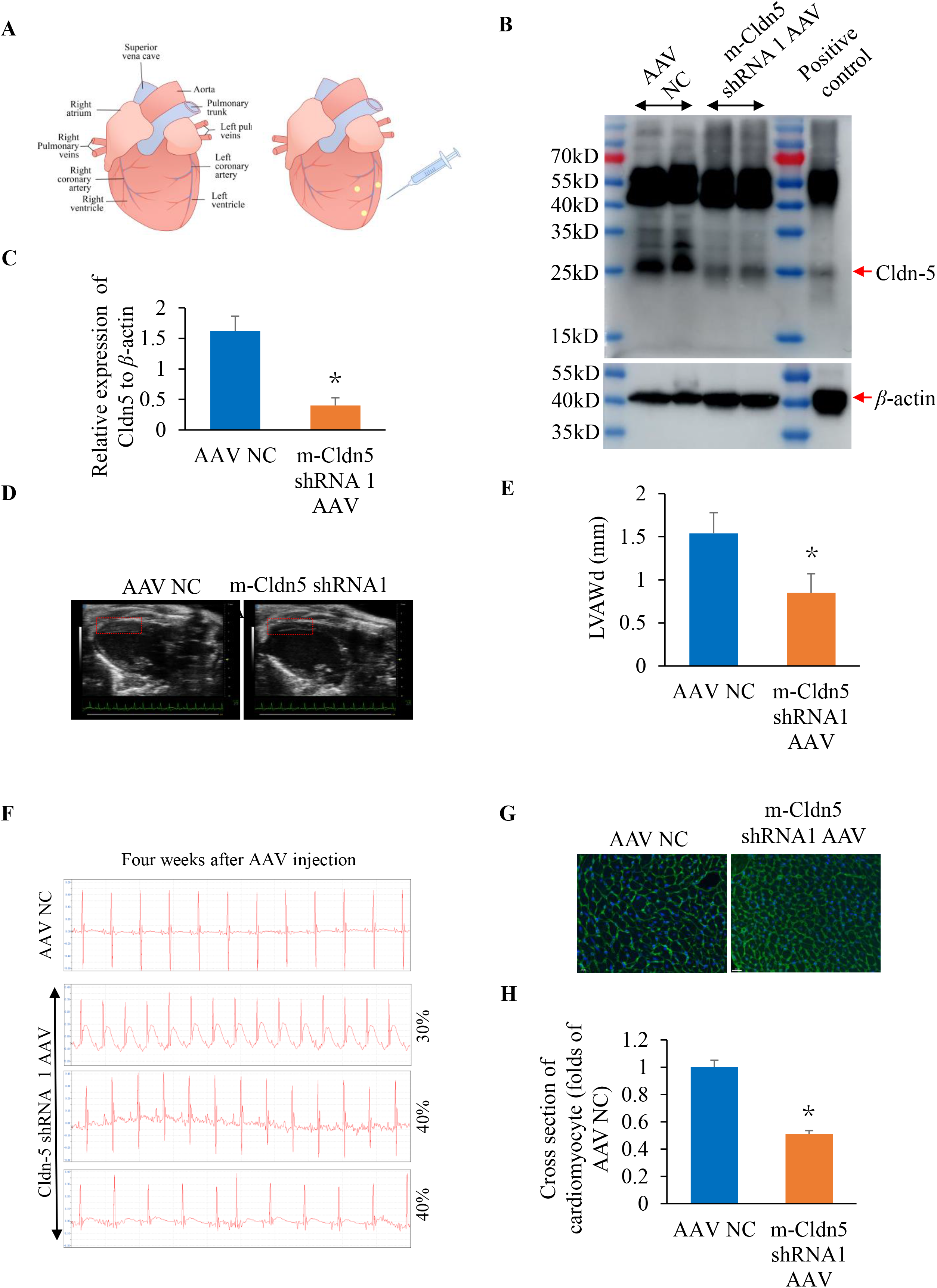
Reduced Cldn5 expression, cardiac atrophy and arrhythmia in mouse heart. (A) Cldn5 shRNA adeno-associated virus (AAV) was injected in three points in the myocardium of the left ventricle (LV). (B) Western blot of claudin-5 in injected area 4 weeks later. Murine lung tissue is used as a positive control. (C) Quantitative analysis of the protein level of claudin-5 in LV myocardium after AAV infection. AAV NC indicates control AAV. * *P* < 0.05 vs. AAC NC; n = 5. (D) Exemplar views of the long axis of LV under echocardiographic observation. A red box indicates the thickness of the LV anterior wall at diastole (LVAWd). (E) Quantitative analysis of the LVAWd. * *P* < 0.05 vs. AAC NC; n = 5. (F) Exemplar images of electrocardiogram (ECG) in mice after 4 weeks of Cldn5 AAV infection in LV myocardium. 30% of mice developed ST elevation; 40% of mice developed AF and 40% of mice developed absence of P-waves prior to QRS. N=10 in each group. (G) The cross-section of cardiomyocytes is stained with WGA after AAV infection for 4 weeks. Scale bar, 20 µm. (H) Quantification of the cross-section of cardiomyocytes. \**P* < 0.05 vs. AAV NC; n = 5.

### Overview of tandem mass tag-based proteomic data

Cardiac tissues from AF and non-AF patients’ left atrial appendages were undertaken proteomic analysis. By strict quality control, we obtained a total of 6,36,881 spectrums (3,05,015 matched), and 50,211 peptides (45,784 unique peptides) were detected among them. Finally, 5,648 proteins were identified, 5,365 of which were quantified (Figure 3A). Biological replicates were validated by the relative standard deviation distribution, which displayed the precision and reproducibility of our proteomic datasets (Figure 3B). Following statistical analysis, 185 proteins with fold-change≥1.50 or ≤ 1/1.5 and *p*-value < 0.05 were considered the DEPs. Among them, a total of 102 up-regulated and 83 down-regulated DEPs were identified when comparing AF with non-AF patients’ heart samples (Figure 3C). These data were clustered on a heatmap to reveal hierarchical commonality in protein abundance for samples within each cardiac tissue type, yet differences in protein abundance when comparing AF and non-AF heart tissues (Figure 3D).

**Fig. 3.**
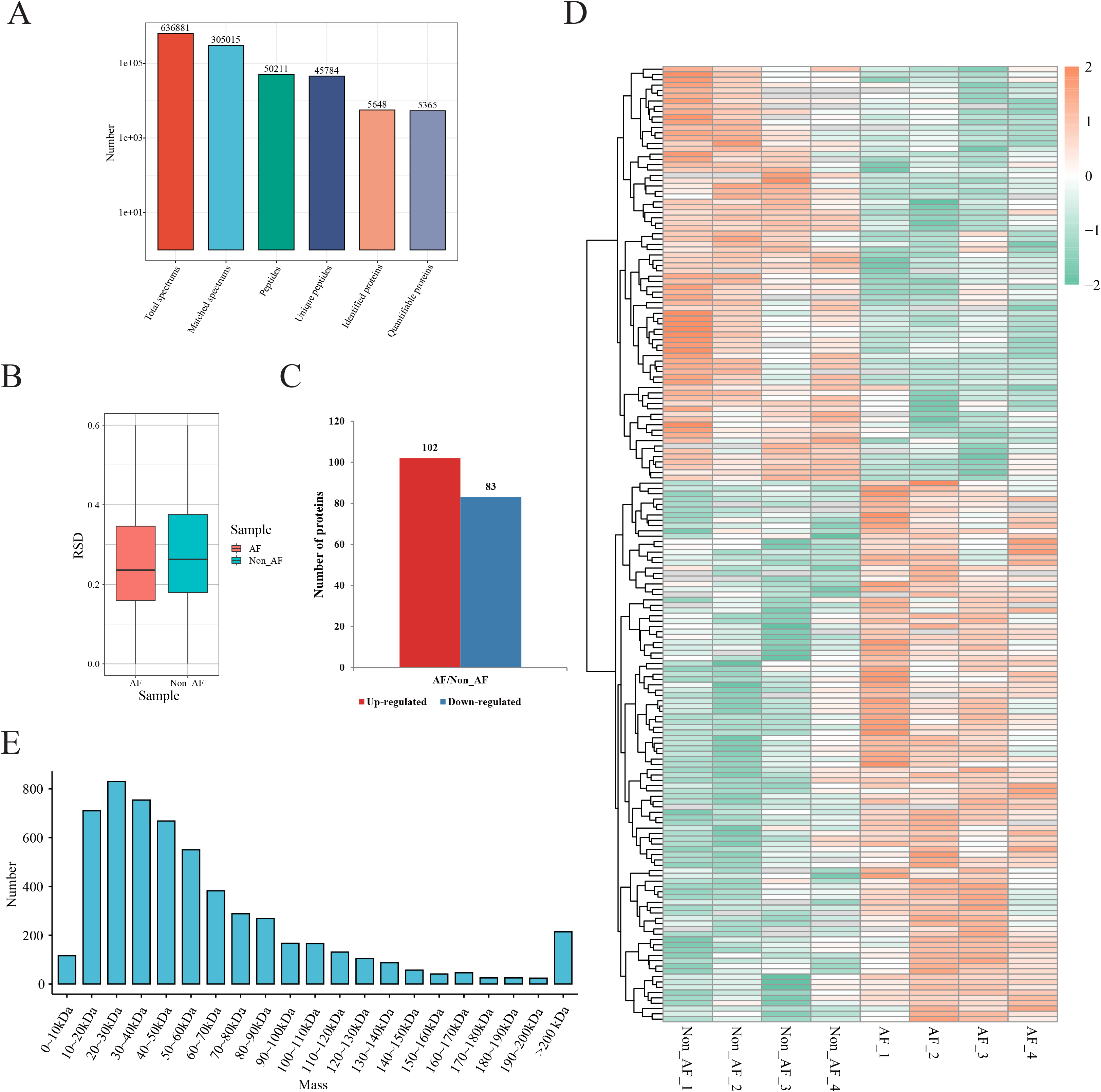
Tandem mass tag (TMT)-based quantitative proteomic sequencing results. (A)Summary of tandem mass spectrometry database search analysis. (B) The relative standard deviation (RSD) of the samples. (C) Statistical analysis of differentially expressed proteins (DEPs). (D) A heatmap with hierarchical clustering of DEPs. (E) The distribution of the molecular size of the identified proteins.

Further analysis showed that the sizes of most proteins were distributed in the range of 10 to 200kDa, which meant reliable results (Figure 3E).

### The cardiac hypertrophy signaling pathway is negatively regulated in AF patients’ left atrium

To confirm the KEGG enrichment analysis, we found the role of DEPs on several signaling pathways especially cardiovascular disease signaling (Figure 4A). We extracted the information solely about cardiovascular disease signaling and found 6 specific cardiovascular signaling pathways, including cardiac hypertrophy signaling (Enhanced), the role of NFAT in cardiac hypertrophy, endothelin-1 signaling, thrombin signaling, HIF1α signaling and Cardiac hypertrophy signaling were negatively regulated by the changed proteins (Figure 4B). Among them, CACNA2D2, CACNB2 and MYL2 were get involved (Figure 4C).

**Fig. 4.**
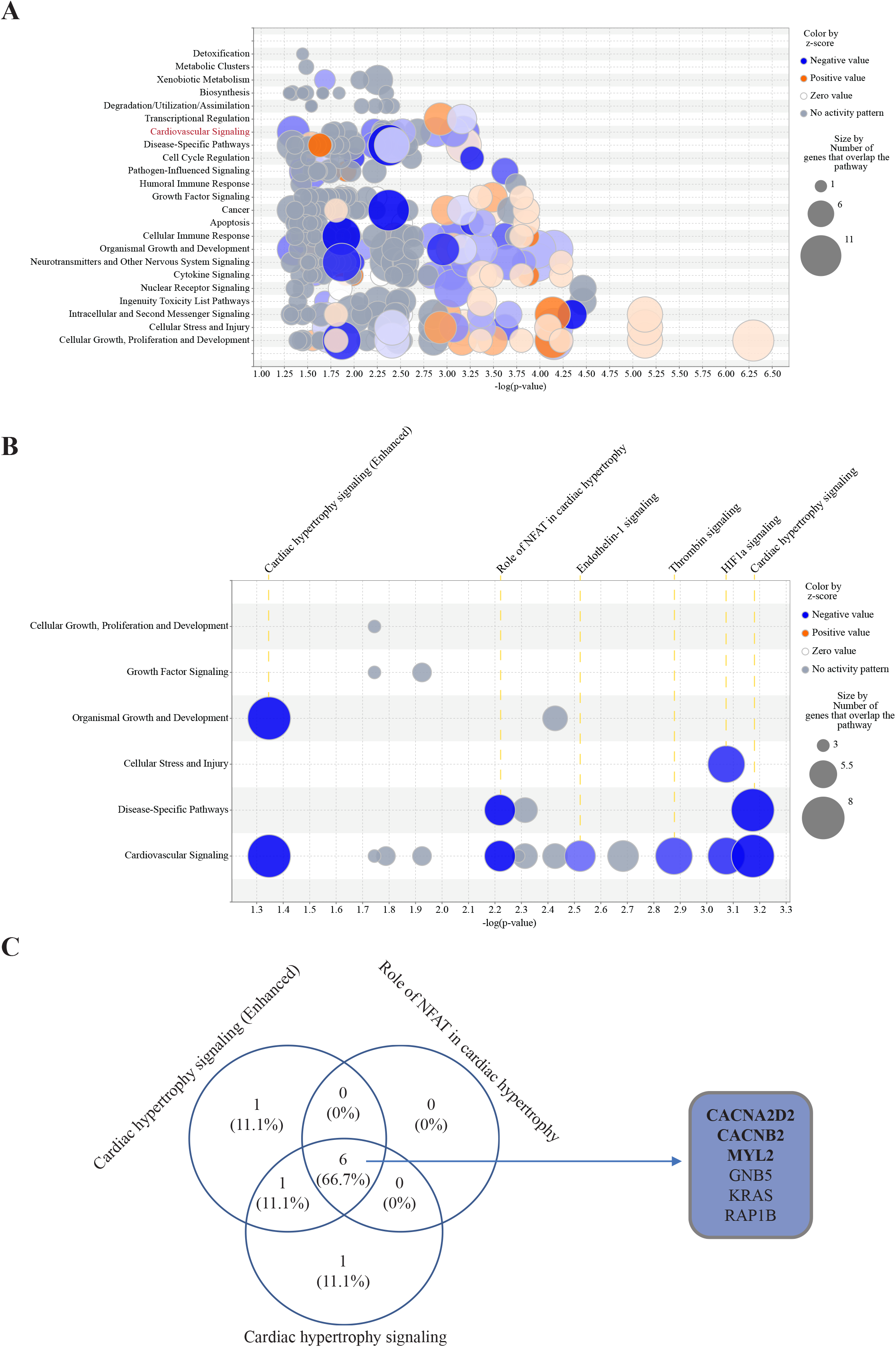
INGENUITY pathway analysis (IPA) results. (A) Bubble chart of metabolic pathway and signal pathway associated with differentially expressed proteins (DEPs). Cardiovascular signaling is marked by the red font. (B) Bubble chart of cardiovascular signaling. (C)Venn diagram demonstrating the number of quantified proteins from the cardiac hypertrophy signaling.

### Hypertrophic and dilated cardiomyopathy KEGG pathways were identified in the AF patients’ cardiomyocyte

To predict the possible roles of DEPs, the INGENUITY pathway analysis was conducted. As shown in Figure 5A, 21 types of cardiovascular diseases were linked to DEPs when compared AF with non-AF atrial tissues. The predictive *p*-value was far away less than 0.05. Among them, abnormal morphology of the heart was associated with 20 changed proteins, abnormality of the heart ventricle was connected with 13 changed proteins and hypertrophic cardiomyopathy was related to 7 changed proteins. The enlarged left atrium was the outcome of mitral stenosis or mitral regurgitation from concentric hypertrophy to eccentric hypertrophy in the development of cardiac remodeling of rheumatic valvular heart disease (Figure 5B). A volcano plot showed the decreased CACNA2D2, CACNB2, MYL2 and MAP6 are highly involved in the development of these three morphological changes of the heart (Figure 5C). Further, KEGG enrichment analysis indicated that these four down-regulated proteins were mainly enriched in the following two pathways: calcium transport and myofibril assembly in cardiomyocytes (Figures 5D and 5E).

**Fig. 5.**
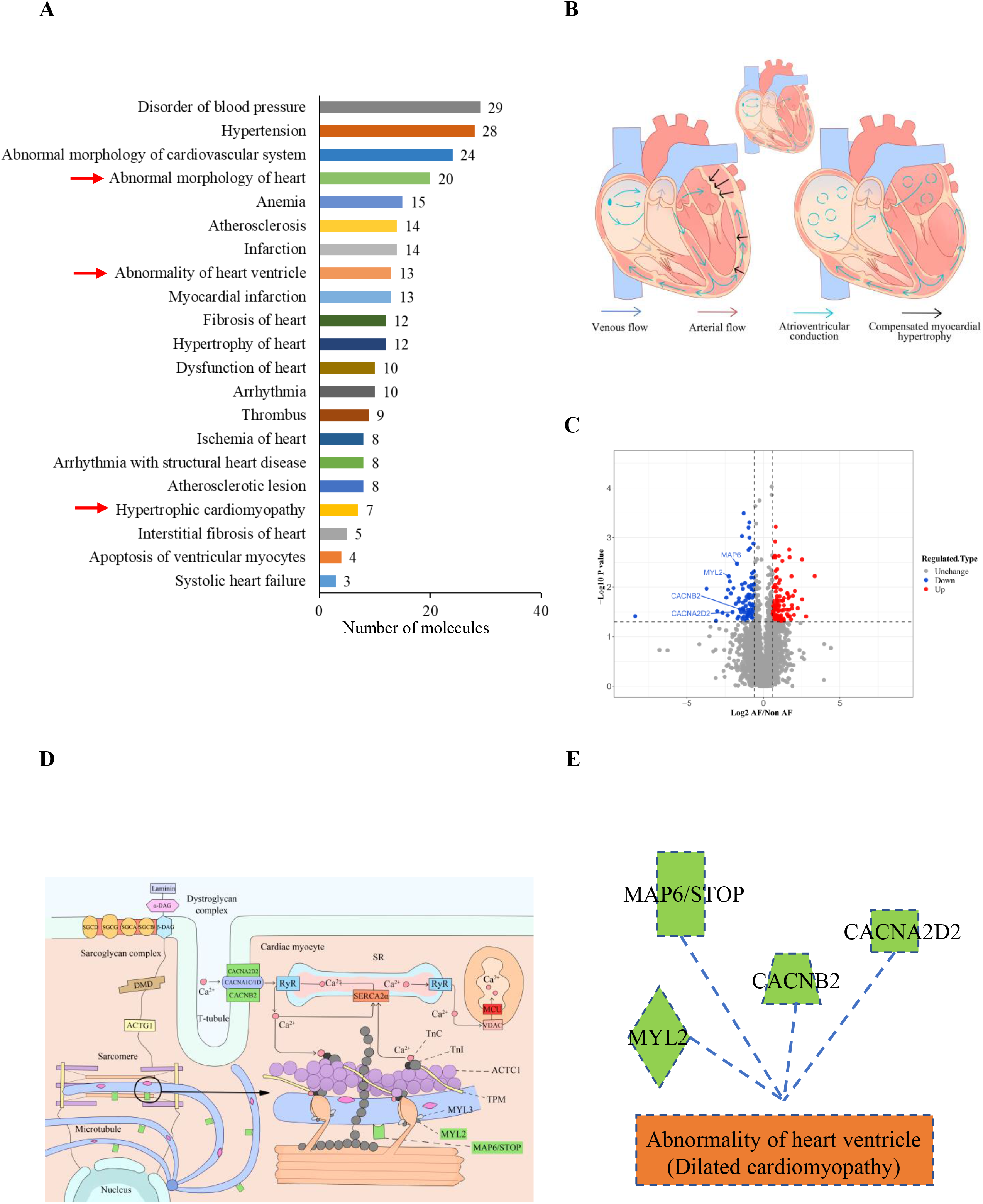
AF patients’ left atrial appendages proteomics exhibits a distinct profile for abnormal heart ventricles. (A) Cardiovascular disease analysis of DAPs. (B) Diagram indicating the transition from hypertrophic cardiomyopathy to dilated cardiomyopathy. (C) Volcano plot of DEPs. Red dots indicate significantly up-regulated proteins, blue dots indicate significant down-regulated proteins and gray dots indicate proteins without differences. Among blue dots, CACNA2D2, CACNB2, MYL2 and MAP6 were highly associated with hypertrophic cardiomyopathy or dilated cardiomyopathy. (D and E) KEGG enrichment analysis of DEPs indicating CACNA2D2, CACNB2, MYL2 and MAP6 were functioned as calcium transport and myofibril assembly. And down-regulation of these proteins will lead to dilated cardiomyopathy.

### Impact of Cldn5 knockdown on dilated cardiomyopathy pathway *in vivo*

The decreased expression of the targeted four DEPs mentioned above in AF patients’ cardiomyocytes was further confirmed by western blot (Figure 6A). Immuno-electron microscopy for Cldn5 showed the morphology of mitochondria collapsed with decreased immunogold-Cldn5 on them (Figure 6B). Similar results were verified in an in-vitro study where Cldn5 was knocked down by shRNA AAV infection for 4 weeks in murine LV. Western blot showed down-regulated protein levels of CACNA2D2, CACNB2, MYL2 and MAP6 after the expression of Cldn5 was disturbed (Figure 6C). The disruption of mitochondrial morphology and distribution as well as depletion of mitochondria were also observed in Cldn5 shRNA AAV-infected myocardium (Figure 6D).

**Fig. 6.**
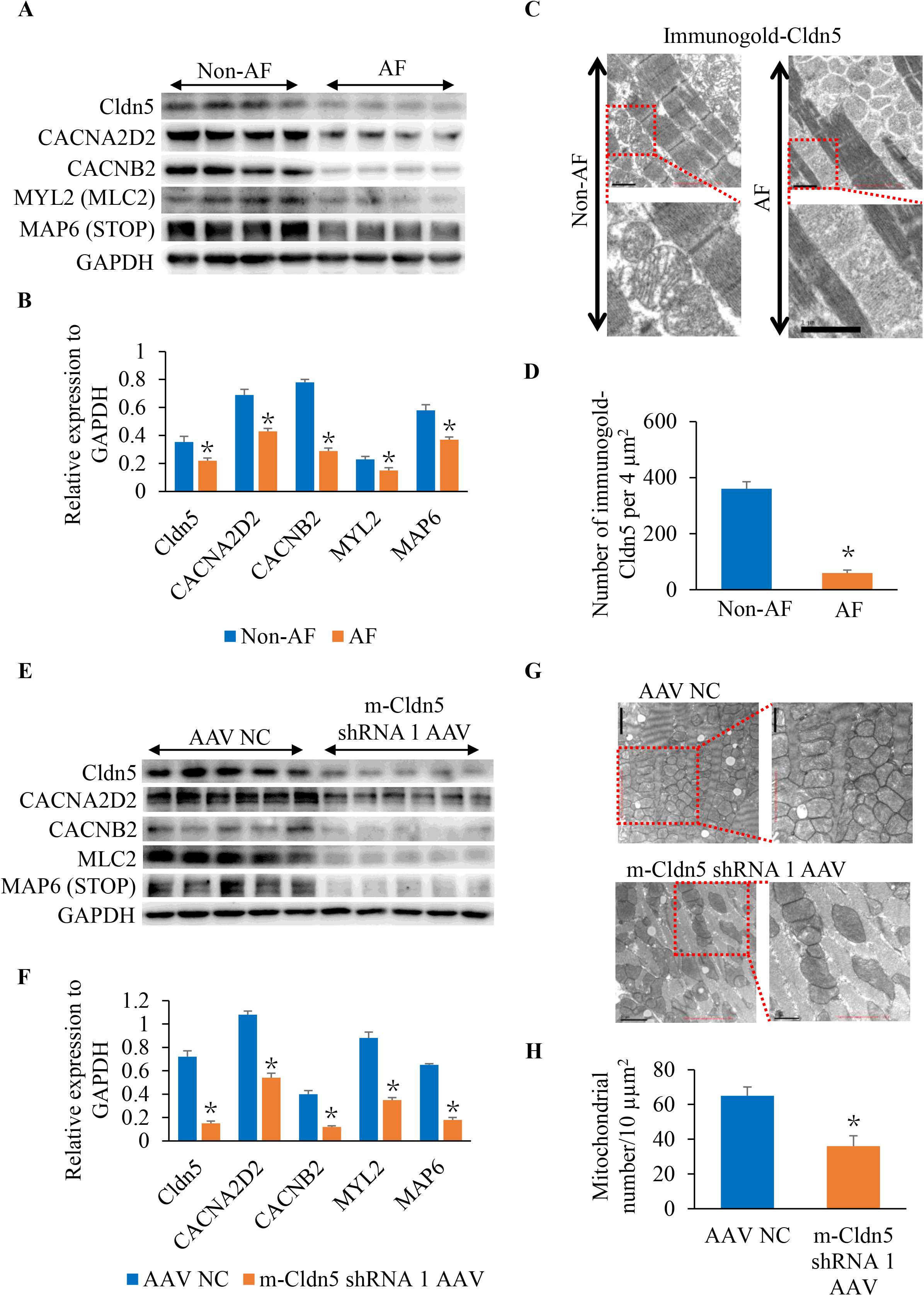
Cldn5 knockdown on dilated cardiomyopathy pathway *in vivo*. (**A and B**) The western blot and quantitative analysis of the Cldn5, CACNA2D2, CACNB2, MYL2 and MAP6 in AF and non-AF left atrial appendages. (C) Representative immunogold-Cldn5 and mitochondrial morphology images were observed under transmission electron microscopy (TEM). Scale bar=1µm. (D) Quantitative analysis of immunogold-Cldn5. (E and F) The western blot and quantitative analysis of the Cldn5, CACNA2D2, CACNB2, MYL2 and MAP6 in the left ventricles of Cldn5 shRNA AAV and AAV NC injected mice. (G) Representative images of mitochondrial morphology under TEM. Scale bar=1µm. (H) Quantitative analysis of mitochondrial number in different groups. \**P* < 0.05 vs. AAV NC; n = 5 in each group.

### Cldn5 knockdown decreases the mitochondrial density and mitochondrial membrane potential *in vitro*

In vitro studies were conducted in HL-1 cells to assess the influence of Cldn5 on the protein levels of CACNA2D2, CACNB2, MYL2 and MAP6 and observe the role of Cldn5 on mitochondrial membrane potential. The four targeted proteins were all down-regulated by si- Cldn5 transfection for 48 hours in the culture medium (Figures 7A and 7B). Decreased number of mitochondria was also observed by Mitotracker staining under confocal microscopy (Figures 7C and 7D). We further detected significantly enhanced fluorescence of JC-1 monomers (green color) after Cldn5 knockdown, which indicated mitochondrial membrane potential was decreased (Figures 7E and 7F).

**Fig. 7.**
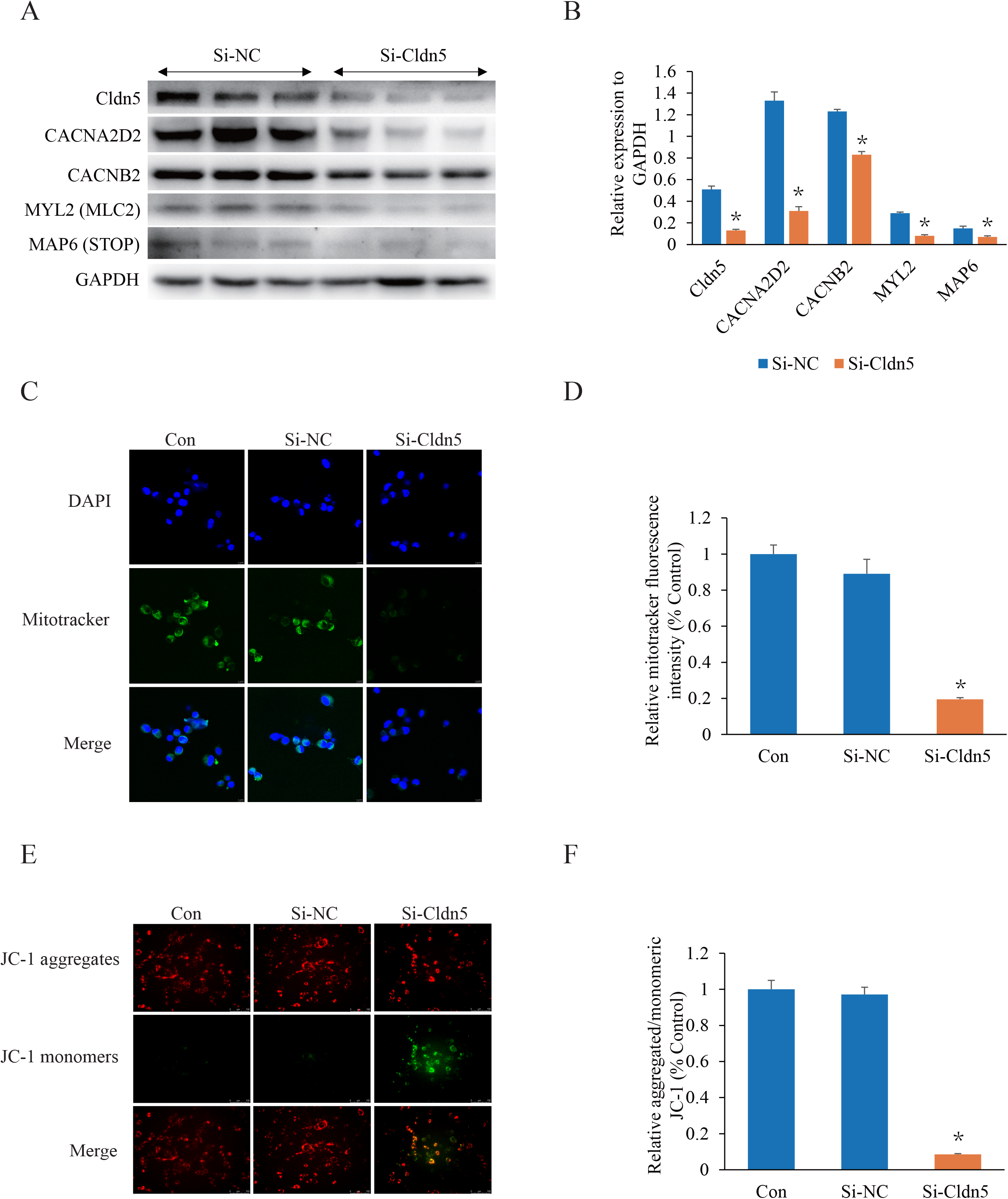
Cldn5 knockdown on the mitochondrial number and membrane potential *in vitro*. (**A and B**) The western blot and quantitative analysis of the Cldn5, CACNA2D2, CACNB2, MYL2 and MAP6 in HL-1 cells after si-Cldn5 transfection for 48 hours. Si-NC is used as the control siRNA. (C) Representative images of mitochondria in HL-1 cells stained with Mitotracker Green. DAPI is used to stain the nucleus. (D) Quantitative analysis of Mitotracker fluorescence intensity. Scale bar=100 µm. (E) Mitochondrial membrane potential was analyzed by JC-1 staining. Scale bar=100 µm. (F) Quantitative analysis of the fluorescence intensity ratio between the red aggregates and green monomer fluorescence. \**P* < 0.05 vs. Si-NC; n = 5 in each group.

## Discussion

### Major findings

Our study firstly demonstrates that reduced expression of Cldn5 in cardiac tissue along with decreased CACNA2D2, CACNB2, MYL2 and MAP6 might induce disturbance of myofibrils and microtubules as well as dysfunction of calcium transport, which leads to myocardial excitation-contraction coupling disorder followed by atrial atrophy, dilation and AF. These findings summarized in Fig. 8 broaden our comprehension of the effect of cell-cell adhesion on arrhythmia.

**Fig. 8.**
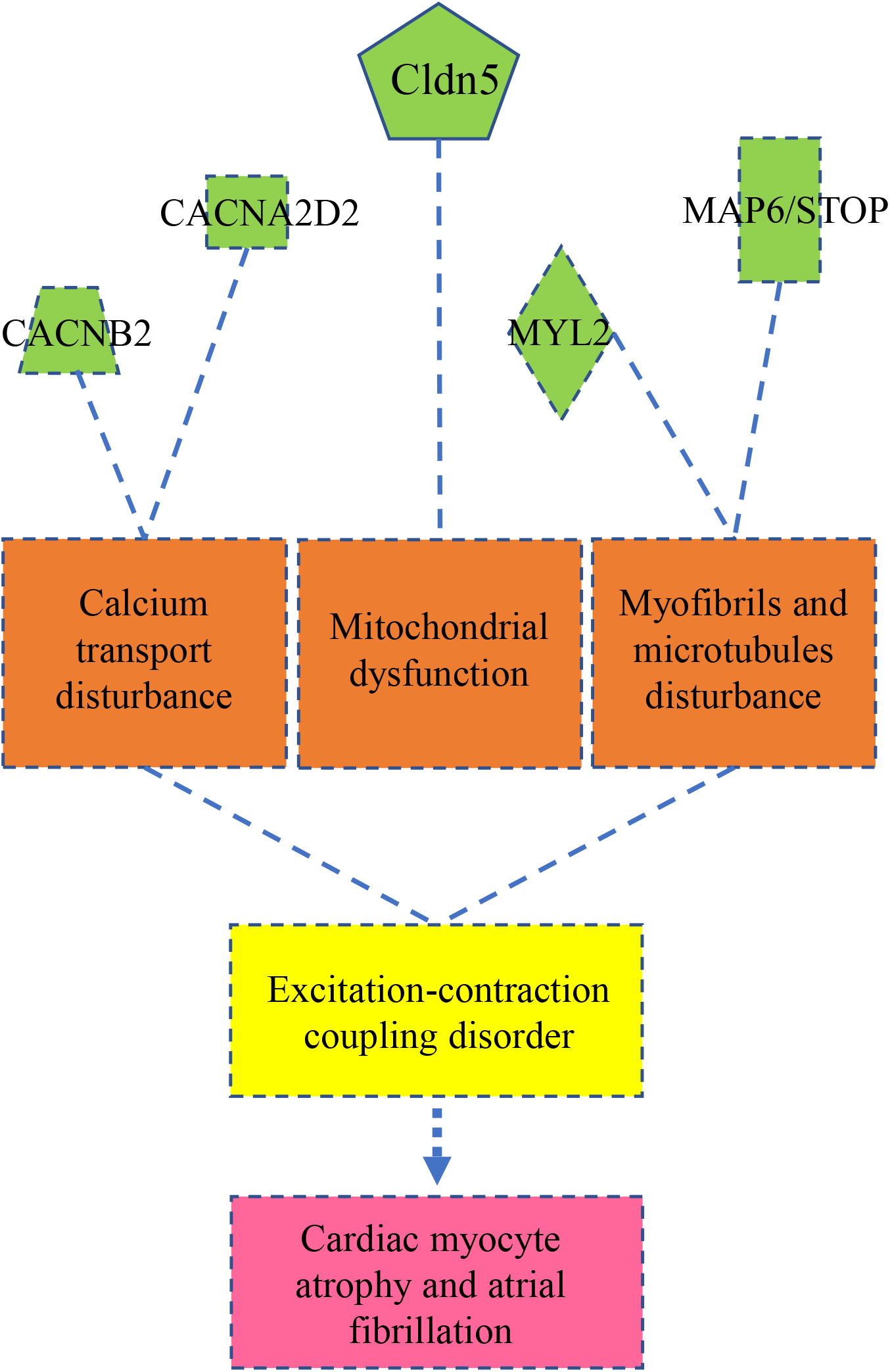
Diagram of claudin-5-mediated signaling. Cldn5 deficiency decreases the expression of CACNA2D2 and CACNB2, leading to calcium transport disturbance. Mitochondrial membrane and membrane potential were decreased after Cldn5 knockdown. Cldn5 deficiency also decreases the expression of MYL2 and MAP6 which are responsible for the disturbance of myofibril assembly and microtubule stabilization. All these effects lead to myocardial excitation-contraction coupling disorder followed by cardiac myocyte atrophy and atrial fibrillation.

### Connexins and AF

The pathogenesis of AF is linked to gap junction remodeling of atrial tissue. Gap junctions are communication structures while tight junctions serve the major functional purpose of providing a “barrier” within the endothelial and epithelial membrane.^14^ Normal cardiac conduction requires gap junction proteins like Cx30, Cx40, Cx43 and Cx45. Since they provide the syncytial properties of the atrium and ventricle, changes in their expression and distribution may lead to arrhythmias. For instance, Cx30 deficient mice had a faster mean daily heart rate than control mice.^15^ Cx30.2 deficient mice also had shorter PQ intervals, elevated AV-nodal conduction velocity and faster ventricular response rates compared to WT littermates.^16^ Cx40 and Cx43 downregulation in the atrium and ventricle of the rabbit’s hearts, respectively, have been thought to enhance the susceptibility to atrial and ventricular tachycardia or fibrillation,^17, 18^ and the heavy ion beam is reportedly promising in the treatment of arrhythmia by increasing the expression of Cx40/43 in animal models.^19^ Additionally, clinical studies also confirmed severely decreased Cx40 levels and reduced overlap with Cx43 in AF patients’ atria.^7^ These lines of evidence have demonstrated the critical role of Cxs in controlling the cardiac rhythm and cardiac conduction, however, the function of the tight junction channel in the pathogenesis of AF is still largely undefined.

### Mechanisms of the causes of Cldn5 deficiency and AF

Cardiomyocyte’s rod-shaped cell morphology is stabilized through specific and direct interaction via the intercalated disk localized at the bipolar ends of the cardiomyocyte, while the lateral cell surface is believed to interact with the extracellular matrix through receptors on the costamere without physical contacts between proximate cardiomyocytes. Cldn5 is found localized in the lateral membranes of cardiomyocytes and is decreased in a mouse model of muscular dystrophy with cardiomyopathy.^20^ Our previous study further found Cldn5 was localized in both sarcolemma and mitochondria of cardiomyocytes and acted as a mitochondrial dynamic regulator preventing mitochondrial fission and preventing the heart from ischemic insult.^13^ However, the electrophysiological role of Cldn5 was still not fully investigated though there was evidence linking tight junction protein to disturbed cardiac rhythm. It has been reported that tight junction protein 1 (TJP1) cardiac-specific deletion transgenic mice developed AV block along with decreased Cx40 expression and intercalated disc localization.^21^ Another study showed that tight junction protein, coxsackievirus-adenovirus receptor (CAR), and heart-specific inducible knockout impaired electrical conduction between the atrium and ventricle in mice.^22^ In the present study, we hypothesized the decreased protein level of Cldn5 in AF patients left atrium was linked to AF, therefore we used Cldn5 shRNA AAV infection to knockdown cardiac Cldn5, and we found ST elevation, ventricular premature beat and AF in Cldn5 ventricle knockdown mice. We also found the expression of Cx40/43 was not changed in western blot after Cldn5 knock down (unpublished data). The implication of these novel findings is that myocardial tight junction protein, Cldn5, maybe also crucial in maintaining normal electrophysiology.

The mechanism underlying the pathogenesis of AF and the other cardiac rhythm disturbance in mice after Cldn5 knockdown in the left ventricle might be different from that of reduced Cldn5 in AF patients’ left atrium though, we captured the common changes of molecules from western blot analysis which showed the protein levels of CACNA2D2, CACNB2, MAP6 and MYL2 were significantly downregulated. Actually, CACNA2D2 serves in vivo as a component of a P/Q-type calcium channel, a functional auxiliary subunit of voltage-gated Ca^2+^ channels, and is indispensable for central nervous system function.^23^ Evidence showed that inhibition or knockdown of CACNA2D2 could induce arrhythmias in rat ischemic hearts.^24^ Moreover, a loss-of-function mutation in another L-type calcium channel subunit gene CACNB2 also has been reported to cause a short QT syndrome subtype.^25^ A missense variant in CACNB2 is also associated with ventricular fibrillation.^26^ Therefore, the downregulated expression of CACNA2D2 and CACNB2 in AF patients’ cardiomyocytes and in Cldn5 knockdown murine cardiomyocytes in the present study suggest a novel role of Cldn5 in modulating calcium transport via voltage-gated Ca^2+^ channels.

The reduction of protein level of Cldn5 is observed in human heart samples from end-stage cardiomyopathy,^11^ and Cldn5 mRNA and protein levels are also specifically reduced in the heart from a mouse model of muscular dystrophy with cardiomyopathy induced by utrophin/dystrophin double knockout.^20^ On the contrary, the over-expression of Cldn5 via adeno-associated virus (AAV) in these double-knockout mice could prevent the development of cardiomyopathy and improve cardiac damage.^27^ Although the connection between Cldn5 and dystrophy in cardiomyopathy or muscular dystrophy is inconclusive, the relation between decreased Cldn5 and cardiomyopathy is apparent. Here we show that significant cardiac myocyte atrophy is common in cardiomyocytes from AF patients’ left atrial appendage and from mouse left ventricle after Cldn5 knockdown. These observations are consistent with the data above suggesting the role of Cldn5 in maintaining the cellular morphology of cardiomyocytes. The mechanism underlying this phenomenon might be associated with decreased MAP6 and MYL2 predicted by proteomic analysis and confirmed by western blot. Since the evidence showed that deletion of the MAP6 resulted in skeletal muscle atrophy and weakness in mice,^28^ and mutation in MYL2 gene was identified in hypertrophic cardiomyopathy (HCM),^29, 30^ we believe that the deficiency of these two proteins after Cldn5 depletion is associated with cardiac myocyte atrophy in the current study.

Furthermore, in our cellular study knockdown Cldn5 in HL-1 cells, we also observed similar results from the in vivo study. Mitochondrial dysfunction might be the key process underlying Cldn5 deficiency-induced arrhythmia since mitochondrial number and membrane potential were decreased after Cldn5 knockdown. These results were consistent with our previous observation that mitochondrial fission was enhanced in cardiomyocytes post hypoxia-reoxygenation when Cldn5 was knocked down by siRNA.^13^ Furthermore, Cldn5 overexpression attenuated myocardial oxidative stress and mitochondrial dysfunction in mice subjected to myocardial ischemia reperfusion injury.^31^ Together these findings suggest that mitochondrial dysfunction induced by the downregulation of Cldn5 along with the significant decrease of CACNA2D2, CACNB2, MAP6 and MYL2 contributed to calcium transport disturbance and myofibrils and microtubules disturbance, leading to myocardial excitation-contraction coupling disorder followed by cardiac myocyte atrophy and fibrillation (Figure 8).

### Study limitations

Although the expression of Cldn5 was decreased in the left atrial appendages in patients with AF, Cldn5 shRNA AAV was injected into the myocardium of the anterior left ventricle rather than the left atrium in this study, suggesting the uncertainly of the areas of Cldn5 expression of the role of AF. Secondly, due to the embryo-lethal, this was a study performed without a Cldn5 gene knockout mouse. An inducible cardiomyocyte-specific Cldn5 deletion mouse (Cldn5^fl/fl^; Myh6^Cre/Esr1*^) will be used in future studies. Moreover, the mechanisms by which Cldn5 knockdown leads to the downregulation of CACNA2D2, CACNB2, MYL2 and MAP6 are not investigated. The interactions between Cldn5 and these four proteins should be considered due to the fact that in failing heart, Cldn5 in cardiomyocytes is reduced and Ephrin-B1 localization is altered,^12^ which hinting that Cldn5 may be required for stabilizing the localization of the other receptors or proteins.

## Conclusions

This is the first clinical study to show the role of Cldn5 on cardiac rhythm and cardiac hypertrophy. The enlargement of LA in Af patients is accompanied by cardiac atrophy and decreased Cldn5 meanwhile cardiac Cldn5 deficiency leads to disturbance of cardiac rhythm and myocyte atrophy. Cell culture findings show decreased mitochondrial numbers and membrane potential. Proteomic study and western blots results demonstrate the detrimental effect of Cldn5 deficiency is associated with the downregulation of CACNA2D2, CACNB2, MYL2 and MAP6. These findings may be considered for the development of promising cardioprotective therapeutics based on claudin-5 to maintain healthy cardiac rhythm and morphology.

## Data Availability

The data that support the findings of this study are available on request from the corresponding author, Dr.Tao Luo?upon reasonable request.

## Acknowledgments

The authors thank Hangzhou Jingjie Biotechnology in Hangzhou, China, for support with proteomics procedures; Qiang Feng from Guangzhou Huayin Health Medical Group Co., Ltd. for excellent technical assistance for transmission electron microscope.

## Sources of Funding

This work was supported by two grants from the National Natural Science Foundation of China (grant no. 82270283 and no. 82060045 to Dr. Tao Luo). A Funded Project from Zunyi Medical University (grant no. [2018]5772-025, to Dr. Tao Luo) and a grant from Guizhou province of China (grant no. [2021]224, to Dr. Tao Luo).

## Disclosures

The authors have no conflicts of interest to disclose.

## Notes

### Competing Interest Statement

The authors have declared no competing interest.

### Clinical Trial

This is not a clinical trial. This is a basic study.

### Author Declarations

Ethical Committee of Zunyi Medical University

## Reference

1. Bosch NA, Cimini J, Walkey AJ. Atrial fibrillation in the icu. Chest. 2018;154:1424-1434

2. Benjamin EJ, Muntner P, Alonso A, Bittencourt MS, Callaway CW, Carson AP, Chamberlain AM, Chang AR, Cheng S, Das SR, Delling FN, Djousse L, Elkind MSV, Ferguson JF, Fornage M, Jordan LC, Khan SS, Kissela BM, Knutson KL, Kwan TW, Lackland DT, Lewis TT, Lichtman JH, Longenecker CT, Loop MS, Lutsey PL, Martin SS, Matsushita K, Moran AE, Mussolino ME, O’Flaherty M, Pandey A, Perak AM, Rosamond WD, Roth GA, Sampson UKA, Satou GM, Schroeder EB, Shah SH, Spartano NL, Stokes A, Tirschwell DL, Tsao CW, Turakhia MP, VanWagner LB, Wilkins JT, Wong SS, Virani SS, American Heart Association Council on E, Prevention Statistics C, Stroke Statistics S. Heart disease and stroke statistics-2019 update: A report from the american heart association. Circulation. 2019;139:e56-e528

3. Wijesurendra RS, Casadei B. Mechanisms of atrial fibrillation. Heart. 2019;105:1860–1867

4. Andersen JH, Andreasen L, Olesen MS. Atrial fibrillation-a complex polygenetic disease. European journal of human genetics : EJHG. 2021;29:1051–1060

5. Guo YH, Yang YQ. Atrial fibrillation: Focus on myocardial connexins and gap junctions. Biology. 2022;11

6. Polontchouk L, Haefliger JA, Ebelt B, Schaefer T, Stuhlmann D, Mehlhorn U, Kuhn-Regnier F, De Vivie ER, Dhein S. Effects of chronic atrial fibrillation on gap junction distribution in human and rat atria. Journal of the American College of Cardiology. 2001;38:883-891

7. Gemel J, Levy AE, Simon AR, Bennett KB, Ai X, Akhter S, Beyer EC. Connexin40 abnormalities and atrial fibrillation in the human heart. Journal of molecular and cellular cardiology. 2014;76:159- 168

8. Patel D, Gemel J, Xu Q, Simon AR, Lin X, Matiukas A, Beyer EC, Veenstra RD. Atrial fibrillation-associated connexin40 mutants make hemichannels and synergistically form gap junction channels with novel properties. FEBS letters. 2014;588:1458–1464

9. Sun Y, Tong X, Chen H, Huang T, Shao Q, Huang W, Laird DW, Bai D. An atrial-fibrillation-linked connexin40 mutant is retained in the endoplasmic reticulum and impairs the function of atrial gap-junction channels. Disease models & mechanisms. 2014;7:561-569

10. Hashimoto Y, Campbell M, Tachibana K, Okada Y, Kondoh M. Claudin-5: A pharmacological target to modify the permeability of the blood-brain barrier. Biological & pharmaceutical bulletin. 2021;44:1380–1390

11. Mays TA, Binkley PF, Lesinski A, Doshi AA, Quaile MP, Margulies KB, Janssen PM, Rafael-Fortney JA. Claudin-5 levels are reduced in human end-stage cardiomyopathy. Journal of molecular and cellular cardiology. 2008;45:81-87

12. Swager SA, Delfin DA, Rastogi N, Wang H, Canan BD, Fedorov VV, Mohler PJ, Kilic A, Higgins RSD, Ziolo MT, Janssen PML, Rafael-Fortney JA. Claudin-5 levels are reduced from multiple cell types in human failing hearts and are associated with mislocalization of ephrin-b1. Cardiovascular pathology : the official journal of the Society for Cardiovascular Pathology. 2015;24:160-167

13. Luo T, Liu H, Chen B, Liu H, Abdel-Latif A, Kitakaze M, Wang X, Wu Y, Chou D, Kim JK. A novel role of claudin-5 in prevention of mitochondrial fission against ischemic/hypoxic stress in cardiomyocytes. The Canadian journal of cardiology. 2021;37:1593–1606

14. Bazzoni G, Dejana E. Endothelial cell-to-cell junctions: Molecular organization and role in vascular homeostasis. Physiological reviews. 2004;84:869–901

15. Gros D, Theveniau-Ruissy M, Bernard M, Calmels T, Kober F, Sohl G, Willecke K, Nargeot J, Jongsma HJ, Mangoni ME. Connexin 30 is expressed in the mouse sino-atrial node and modulates heart rate. Cardiovascular research. 2010;85:45-55

16. Kreuzberg MM, Schrickel JW, Ghanem A, Kim JS, Degen J, Janssen-Bienhold U, Lewalter T, Tiemann K, Willecke K. Connexin30.2 containing gap junction channels decelerate impulse propagation through the atrioventricular node. Proceedings of the National Academy of Sciences of the United States of America. 2006;103:5959–5964

17. Amino M, Yamazaki M, Yoshioka K, Kawabe N, Tanaka S, Shimokawa T, Niwa R, Tomii N, Kabuki S, Kunieda E, Yagishita A, Ikari Y, Kodama I. Heavy ion irradiation reduces vulnerability to atrial tachyarrhythmias - gap junction and sympathetic neural remodeling. Circulation journal : official journal of the Japanese Circulation Society. 2023;87:1016–1026

18. Amino M, Yoshioka K, Tanabe T, Tanaka E, Mori H, Furusawa Y, Zareba W, Yamazaki M, Nakagawa H, Honjo H, Yasui K, Kamiya K, Kodama I. Heavy ion radiation up-regulates cx43 and ameliorates arrhythmogenic substrates in hearts after myocardial infarction. Cardiovascular research. 2006;72:412-421

19. Amino M, Yoshioka K, Kamada T, Furusawa Y. The potential application of heavy ion beams in the treatment of arrhythmia: The role of radiation-induced modulation of connexin43 and the sympathetic nervous system. International journal of particle therapy. 2018;5:140-150

20. Sanford JL, Edwards JD, Mays TA, Gong B, Merriam AP, Rafael-Fortney JA. Claudin-5 localizes to the lateral membranes of cardiomyocytes and is altered in utrophin/dystrophin-deficient cardiomyopathic mice. Journal of molecular and cellular cardiology. 2005;38:323-332

21. Dai W, Nadadur RD, Brennan JA, Smith HL, Shen KM, Gadek M, Laforest B, Wang M, Gemel J, Li Y, Zhang J, Ziman BD, Yan J, Ai X, Beyer EC, Lakata EG, Kasthuri N, Efimov IR, Broman MT, Moskowitz IP, Shen L, Weber CR. Zo-1 regulates intercalated disc composition and atrioventricular node conduction. Circulation research. 2020;127:e28-e43

22. Lisewski U, Shi Y, Wrackmeyer U, Fischer R, Chen C, Schirdewan A, Juttner R, Rathjen F, Poller W, Radke MH, Gotthardt M. The tight junction protein car regulates cardiac conduction and cell-cell communication. The Journal of experimental medicine. 2008;205:2369–2379

23. Ivanov SV, Ward JM, Tessarollo L, McAreavey D, Sachdev V, Fananapazir L, Banks MK, Morris N, Djurickovic D, Devor-Henneman DE, Wei MH, Alvord GW, Gao B, Richardson JA, Minna JD, Rogawski MA, Lerman MI. Cerebellar ataxia, seizures, premature death, and cardiac abnormalities in mice with targeted disruption of the cacna2d2 gene. The American journal of pathology. 2004;165:1007–1018

24. Zhang J, Wu L, Li Z, Fu G. Mir-1231 exacerbates arrhythmia by targeting calciumchannel gene cacna2d2 in myocardial infarction. American journal of translational research. 2017;9:1822-1833

25. Zhong R, Zhang F, Yang Z, Li Y, Xu Q, Lan H, Cyganek L, El-Battrawy I, Zhou X, Akin I, Borggrefe M. Epigenetic mechanism of l-type calcium channel beta-subunit downregulation in short qt human induced pluripotent stem cell-derived cardiomyocytes with cacnb2 mutation. Europace : European pacing, arrhythmias, and cardiac electrophysiology : journal of the working groups on cardiac pacing, arrhythmias, and cardiac cellular electrophysiology of the European Society of Cardiology. 2022;24:2028–2036

26. Zhong R, Schimanski T, Zhang F, Lan H, Hohn A, Xu Q, Huang M, Liao Z, Qiao L, Yang Z, Li Y, Zhao Z, Li X, Rose L, Albers S, Maywald L, Muller J, Dinkel H, Saguner A, Janssen JWG, Swamy N, Xi Y, Lang S, Kleinsorge M, Duru F, Zhou X, Diecke S, Cyganek L, Akin I, El-Battrawy I. A preclinical study on brugada syndrome with a cacnb2 variant using human cardiomyocytes from induced pluripotent stem cells. International journal of molecular sciences. 2022;23

27. Delfin DA, Xu Y, Schill KE, Mays TA, Canan BD, Zang KE, Barnum JA, Janssen PM, Rafael-Fortney JA. Sustaining cardiac claudin-5 levels prevents functional hallmarks of cardiomyopathy in a muscular dystrophy mouse model. Molecular therapy : the journal of the American Society of Gene Therapy. 2012;20:1378–1383

28. Sebastien M, Giannesini B, Aubin P, Brocard J, Chivet M, Pietrangelo L, Boncompagni S, Bosc C, Brocard J, Rendu J, Gory-Faure S, Andrieux A, Fourest-Lieuvin A, Faure J, Marty I. Deletion of the microtubule-associated protein 6 (map6) results in skeletal muscle dysfunction. Skeletal muscle. 2018;8:30

29. Yin K, Ma Y, Cui H, Sun Y, Han B, Liu X, Zhao K, Li W, Wang J, Wang H, Wang S, Zhou Z. The co- segregation of the myl2 r58q mutation in chinese hypertrophic cardiomyopathy family and its pathological effect on cardiomyopathy disarray. Molecular genetics and genomics : MGG. 2019;294:1241–1249

30. Richard P, Charron P, Carrier L, Ledeuil C, Cheav T, Pichereau C, Benaiche A, Isnard R, Dubourg O, Burban M, Gueffet JP, Millaire A, Desnos M, Schwartz K, Hainque B, Komajda M, Project EHF. Hypertrophic cardiomyopathy: Distribution of disease genes, spectrum of mutations, and implications for a molecular diagnosis strategy. Circulation. 2003;107:2227-2232

31. Jiang S, Liu S, Hou Y, Lu C, Yang W, Ji T, Yang Y, Yu Z, Jin Z. Cardiac-specific overexpression of claudin-5 exerts protection against myocardial ischemia and reperfusion injury. Biochimica et biophysica acta. Molecular basis of disease. 2022;1868:166535

